# Detection of SARS-CoV-2 RNA in fourteen wastewater treatment systems in Uttarakhand and Rajasthan States of North India

**DOI:** 10.1101/2020.09.18.20197178

**Authors:** Sudipti Arora, Aditi Nag, Ankur Rajpal, Satya Brat Tiwari, Jasmine Sethi, Devanshi Sutaria, Jayana Rajvanshi, Sonika Saxena, Sandeep Srivastava, A. A. Kazmi, Vinay Kumar Tyagi

## Abstract

We investigated the presence of Severe Acute Respiratory Syndrome Coronavirus 2 (SARS-CoV-2) RNA at different treatment stages of 15 wastewater treatment plants (WWTPs) in two North Indian states of Rajasthan and Uttarakhand. Untreated (influent), biologically treated, and disinfected wastewater samples were collected from May to August 2020. The qualitative analysis of the wastewater for the presence of SARS-CoV-2 RNA was done using different pre-processing methods. SARS-CoV-2 RNA was detected in 11 out of 39 wastewater samples in Jaipur district and 5 out of 17 wastewater samples in Haridwar District using Reverse-Transcriptase Quantitative Polymerase Chain Reaction (RT-qPCR) for qualitative detection. None of the 56 samples tested for post-secondary or tertiary treatment were found positive for SARS-CoV-2 RNA. The findings indicate that there are no SARS-CoV-2 related risks involved with using the treated effluent for non-potable applications. In contrast, untreated wastewater may be a potential route of viral transmission to the WWTP and sanitation workers. Future studies are imperative to understand the survival rates of these viruses in wastewater.

**Graphical Abstract:** 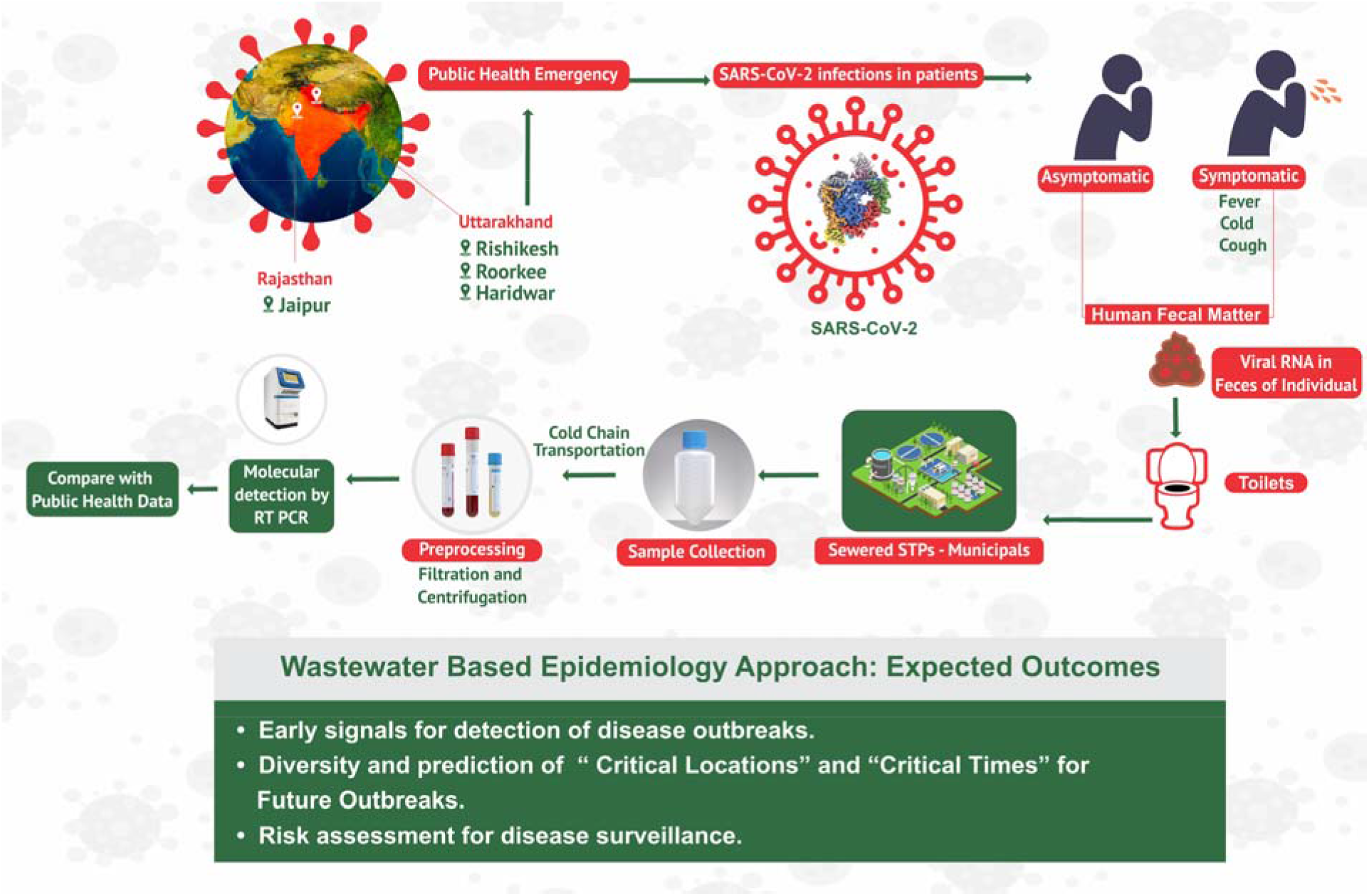

**Highlights:**

- Mild to moderate genome load observed in the municipal wastewater samples.
- Increased patient numbers post-lockdown correspond to a decrease in the C_T_ value of genes.
- Presence of SARS-CoV-2 genome load was observed in untreated wastewater.
- E gene was present in abundance in wastewaters as compared to the N gene and RdRp gene.
- SARS-CoV-2 genome load was absent in secondary and tertiary treated effluent.

## 1. Introduction

The coronavirus disease (COVID-19) caused by Severe Acute Respiratory Syndrome Coronavirus 2 (SARS-CoV-2) has emerged as a worldwide public health emergency within few months of its outbreak in Wuhan, China. Its spread is confirmed in more than 213 countries/regions worldwide (worldometers.info). COVID-19 is highly contagious and spreads through different routes, like air droplets, and physical contact. Hence, early detection and rapid containment protocols are crucial for its control and elimination. Especially for developing countries, many of whom are densely populated and lack robust public health infrastructure. Relying on clinical testing alone for detection and control is insufficient due to the scale of the spread and many asymptomatic and pauci-asymptomatic cases. Wastewater based epidemiology (WBE) or wastewater surveillance can be a useful tool for community-wide detection as both viable SARS-CoV-2, and viral RNA are shed in bodily excreta, including saliva, sputum, and feces, subsequently disposed of in wastewater.

WBE implies the extraction, detection, analysis and interpretation of the biomarkers from the wastewater (generally obtained from a limited geographic area) (Sims and Hordern, 2020). It has been successfully used to predict disease outbreak previously, like, the spread of wild poliovirus type-1 (WPV-1) in Egypt and Palestine (WHO, 2013). Similarly, SARS-CoV-2 RNA fragments were found in wastewater in Brazil (Fongaro et al., 2020), Italy (La Rosa et al., 2020), the Netherlands (Medema et al., 2020) and Spain (Randazzo et al. (2020) before the first clinically confirmed cases there. Various other studies detected SARS-CoV-2 RNA in wastewater across the globe (Ahmed et al., 2020; La Rosa et al., 2020; Lodder and Husman, 2020; Medema et al., 2020; Rimoldi et al., 2020; Wu et al., 2020; Wurtzer et al., 2020), and wastewater surveillance has been suggested as a non-invasive early-warning tool for monitoring the status and trend of COVID-19 infection and for tuning public health response (Daughton, 2020; Mallapaty, 2020; Naddeo and Liu, 2020). Combinations of clinical and environmental surveillance methods have proven to be useful for planning, implementing and evaluating public health practices (Kroiss et al., 2018). However, current evidence indicates the need for a better understanding of the role of wastewater as potential sources of epidemiological data and as a factor of public health risk. A recent study suggests that SARS-CoV-2 can be infective in aerosols for up to 16 hours (Fears et al., 2020), thus implying possible human health risk due to wastewater aerosolization, especially among sanitation workers. Thus, it is imperative to investigate the presence of viral RNA in both untreated and treated wastewater. Further, there are apprehensions about implementing WBE in developing countries due to poor water supply network and sewerage system. While infrastructure development is quintessential and a long-drawn process, sufficient good quality data on WBE from these regions can be useful for future planning and computational modeling.

Most studies on WBE for SARS-CoV-2 are reported from developed countries. Therefore, this study tries to bridge the crucial information gap regarding WBE in a developing region, specifically India. The objective of this study is to detect SARS-CoV-2 RNA fragments in both untreated and treated wastewater samples collected from multiple locations. Experiments were carried out to detect the presence of SARS-CoV-2 in influent, secondary treated and tertiary treated effluent samples from 15 wastewater treatment systems of four cities (Roorkee, Rishikesh, Haridwar, Jaipur) of two North Indian states of Uttarakhand and Rajasthan, and to possibly decipher the potential of current biological treatment systems for removal of the virus. The samples from multiple locations make the study more representative and indicate the potential application of WBE across diverse climatic conditions. This study will add to the existing literature on WBE and contribute to an efficient and resilient public health emergency response mechanism for the future.

## 2. Materials and methods

### 2.1. Sample collection

Wastewater samples (grab) were collected during nine different time points from seven wastewater treatment facilities of Jaipur city (Rajasthan). Grab, and composite samples were collected during three different time points from eight wastewater treatment facilities at Uttarakhand state (Rishikesh, Haridwar, Roorkee). The sampling was carried out during the months of May to August 2020. Untreated wastewater (n= 23 and 9), secondary treated effluent (n= 5 and 2) and tertiary treated effluent (n= 9 and 6) were collected from Rajasthan and Uttarakhand states, respectively. The locations of the WWTPs are highlighted in Figure 1 and summarized in Table 1. During sampling, proper precaution and safety measures like using standard personal protection equipment (PPE), were followed. Samples were collected in clean pre-sterilized bottles and transported to Dr. B. Lal Clinical Laboratory, Jaipur for pre-processing and RNA extraction.

**Table 1.** Sampling locations in Rajasthan and Uttarakhand states

| S.No. | Sampling Location | Secondary Treatment | Tertiary Treatment | Type of Sample |
| --- | --- | --- | --- | --- |
| **(A) Rajasthan** |  |  |  |  |
| 1 | Rambagh Circle, Jaipur<br>26.8963° N, 75.8100° E | MBBR | UV | Influent, Effluent |
| 2 | Central Park, Jaipur<br>26.9048° N, 75.8073° E | SBR | Cl | Influent, Effluent |
| 3 | Dhelawas, Jaipur<br>27.3735° N, 75.8926° E | ASP | -- | Influent, Effluent |
| 4 | Jawahar Circle, Jaipur<br>26.8414°N, 75.8°E | MBBR | UV | Influent, Secondary treated, Tertiary treated (Disinfection) |
| 5 | Brahmpuri, Jaipur<br>26.9373° N, 75.8250° E | SBR | -- | Influent, Effluent |
| 6 | MNIT, Jaipur<br>26.8640° N, 75.8108° E | MBBR | Cl | Influent, Effluent |
| 7 | Dravyawati River Project, Jaipur<br>26.7980° N, 75.8039° E | SBR | Cl | Influent, Effluent |
| **(B) Uttarakhand** |  |  |  |  |
| 8 | IIT Roorkee<br>29.8649° N, 77.8965° E | SBR | UV | Influent, Secondary treated, Tertiary treated (Disinfection) |
| 9 | Muni ki Reti, Rishikesh<br>30.1199° N, 78.3031° E | MBBR | Cl | Influent, Effluent |
| 10 | Swarg Ashram, Rishikesh<br>30.1165° N, 78.3131° E | SBR | -- | Influent |
| 11 | Chandreshwar Nagar, Rishikesh<br>30.1115° N, 78.3056° E | MBBR | Cl | Influent, Effluent |
| 12 | Sarai, Haridwar<br>29.9043° N, 78.1080° E | SBR | Cl | Influent, Effluent |
| 13 | Jagjeetpur, Haridwar | MBBR | Cl | Influent, Effluent |
|  | 29.9174° N, 78.1316° E |  |  |  |
| 14 | Sewage Pump House,<br>Haridwar<br>29°56'36.00°N,<br>78°9'20.06°E | -- | -- | Influent |

MBBR= Moving bed biofilm reactor, SBR= Sequencing batch reactor, ASP= Activated sludge process, UV= Ultra-violet, Cl= Chlorination

**Fig. 1.**
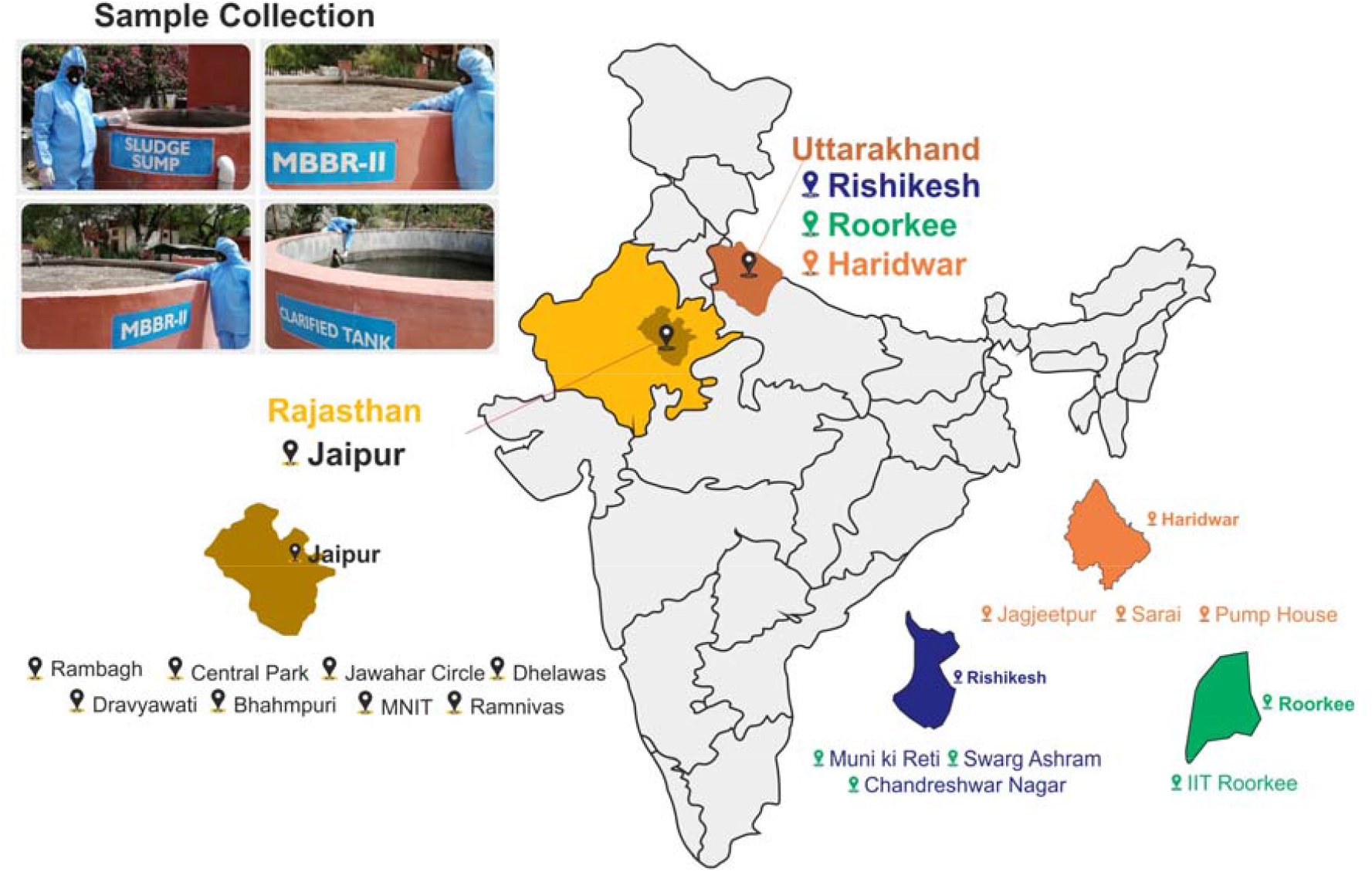
Locations of samples collection in the Uttarakhand and Rajasthan states of India.

### 2.2. Sample pre-processing

The samples were pre-processed using two different methods specified in Figure 2. In method A, a 50 ml sample was transferred into sterile falcon tubes in Biosafety Cabinet (BSL-II) followed by surface sterilization of the falcons using 70% ethanol and exposure to UV light for 30 minutes. The heat inactivation of the virus was then done by placing the falcon tubes in a water bath at 60°C for 90 mins. The samples were further filtered through a 0.45μm membrane using a vacuum filter assembly. The filtrate was then transferred to a fresh falcon containing 4g PEG and 0.9g NaCl. The content was dissolved through manual mixing followed by centrifugation at 4°C for 30mins at 7,000 rpm. The pellet obtained was then resuspended in 1X Phosphate Buffer Saline (PBS).

**Fig. 2.**
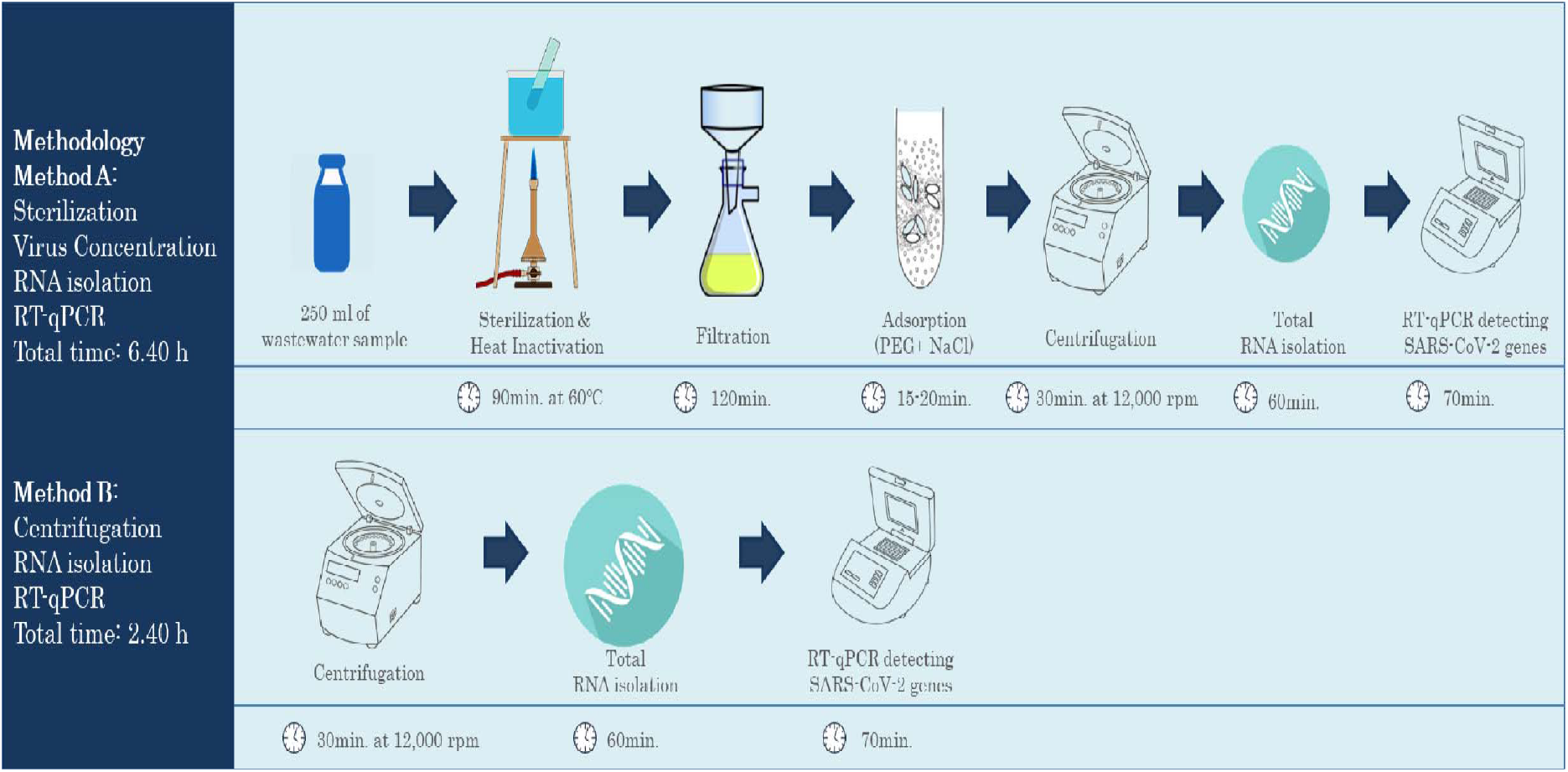
Methodology for the detection of SARS-CoV-2 viral genome through RT-qPCR in wastewater samples

Method B used for the detection of the SARS-CoV-2 virus was performed by the transfer of a 1ml sample in a 1.7ml centrifuge tube followed by centrifugation at 7,000 rpm for 15mins. The supernatant was collected in a fresh tube and was again centrifuged at 7,000 rpm for 15mins. The supernatant thus obtained was used for nucleic acid extraction. Since both methods A and B showed similar patterns of detection of SARS-CoV-2 genes for early time points of the collection time window (Arora et al., 2020); all further samples from July 2020 onwards were pre-processed following method B.

### 2.3. Nucleic acid extraction

The viral RNA molecules present in the wastewater samples from May to early July 2020 were isolated using a Biospin kit (Cat# BSC77M1). As per the vendor’s instructions, 10 μl proteinase K and 200 μl of lysis buffer were added to 200μl of sample into a 1.5 ml centrifuge tube followed by vortex mixing and incubation at 56 °C for 15 minutes in a heating block. 250 μl of ethanol was then added to the sample and mixed by vortexing for 15 secs. The mixture was then transferred to the spin column and centrifuged at 10,000 g followed by sequential washing with the three wash buffers provided in the kit followed by centrifugation at 10,000g for 1 min at each washing step. After complete drying of the spin column, the RNA was eluted out using a 50-100μl elution buffer. centrifugation was done at 12,000 g for 1 minute. The RNA from samples collected in late July to August has been extracted using the automated KingFisher Flex System™. (Cat#5400610).

### 2.4. Method for RT-qPCR based qualitative detection of viral RNA from sewage samples

For the qualitative detection of SARS-CoV-2 in the wastewater samples, RT-PCR was performed using FDA approved Allplex™ 2019-nCoV Assay kit (Cat# RP10244Y, 208 RP10243X) as per protocol standardized in the laboratory (Arora et al., 2020). Recently published Real Time-PCR assays were used as the basis for the detection of SARS-Co-V-2 in wastewater samples (Ahmed et al., 2020; Corman et al., 2020). RT-PCR assays were performed using two of the ICMR approved kits for standardizing detection of viral genome in wastewater samples. The two kits used were FDA approved Allplex™ 2019-nCoV Assay kit (cat# RP10244Y, RP10243X) and TaqPath™ COVID-19 Combo Kit (Cat#A47814) for the qualitative detection of SAR-CoV-2 genomic RNA in the sample on Applied Biosystems™ QuantStudio™ 5. Once the protocol was established and the two kits performed without any noticeable difference in efficiency, further samples were all tested by Allplex™ kit. The protocol for the same is as follows: the mastermix was prepared using the kit content which was composed of Amplification and detection reagent, enzyme mix for one-time RT-PCR, buffer containing dNTPs, buffer for one-step PCR and RNase free water. Each PCR tube contained 8 μl RNA sample, 5μl 2019-nCoV MOM, 5 μl Real-time One-step buffer and 2μl Real-time One-step enzyme and the final volume of the mixture was adjusted to 25μl using RNase free water. A list of different fluorophores used for the detection run is given in Table 2. Thermal cycling reactions were performed at 50 °C for 20 minutes, 95 °C for 15 minutes, 44 cycles at 94 °C for 15 seconds, and 45 cycles at 58 °C for 30 seconds, in a thermal cycler. For each run, a set of positive and negative controls were included.

**Table 2.**
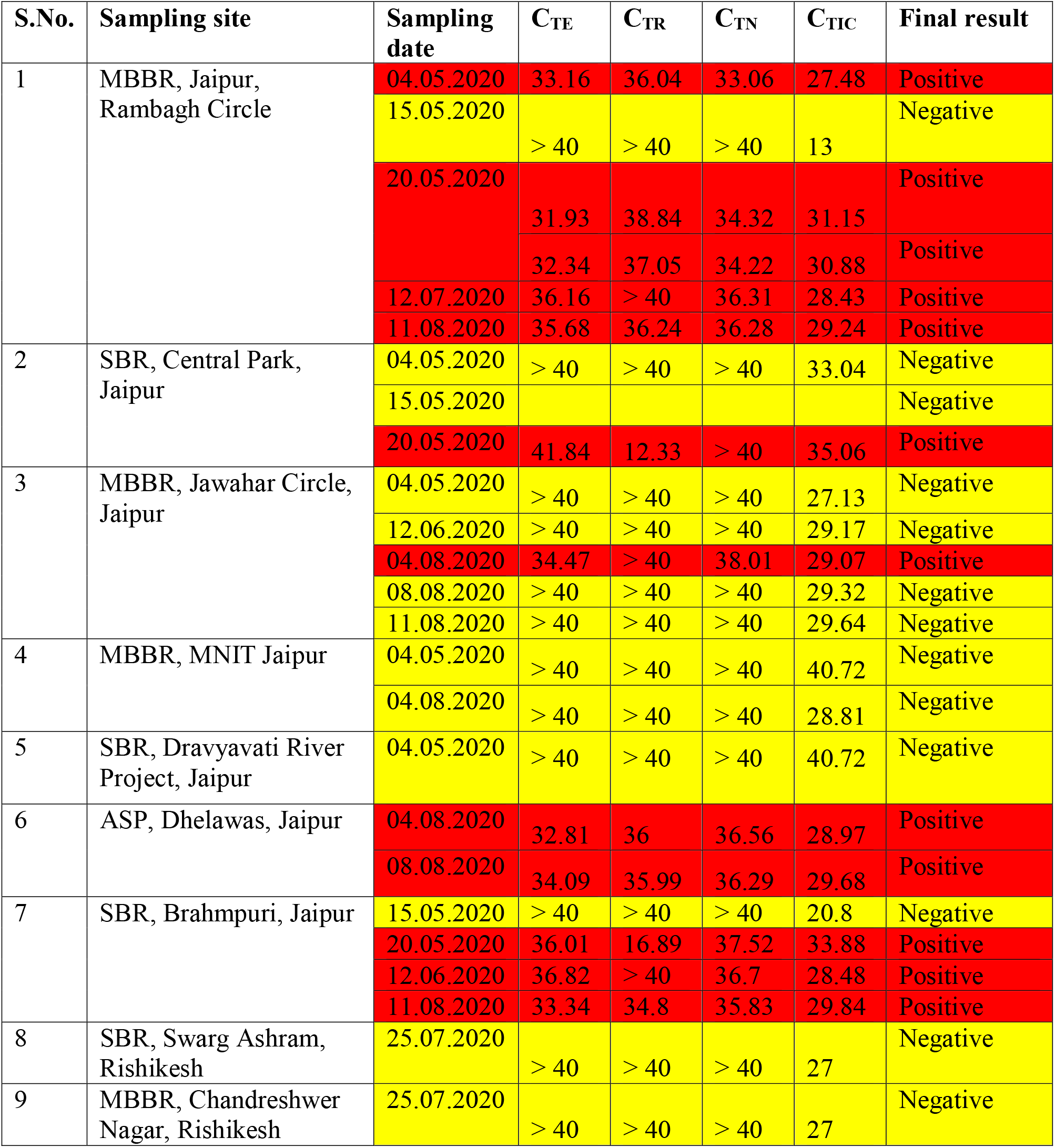
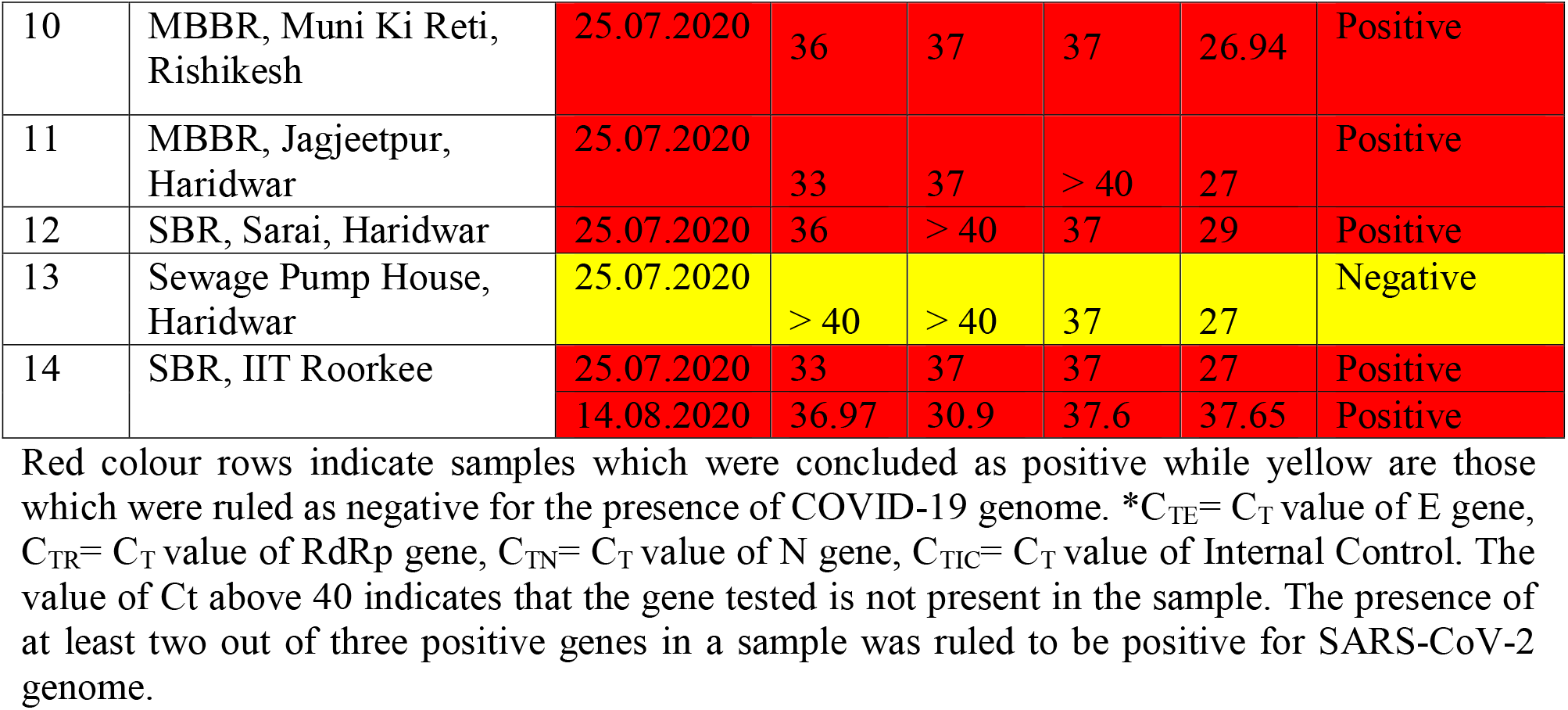
SARS CoV-2 RNA in untreated wastewater samples collected from different WWTPs

## 3. Results and discussion

### 3.1. Detection of SARS-CoV-2 RNA in raw wastewater samples corresponds to epidemiological data for detection of Hotspots

The wastewater influent samples were collected from various sites in Rajasthan and Uttarakhand and tested for the presence of the viral genome. In Rajasthan, the sampling was started in early May 2020. In Uttarakhand, the sampling was started in July 2020. The early sampling in Rajasthan coincided with the duration of community restrictions and lockdown and followed the gradual relaxation in the same (Arora et al. 2020). The sampling was carried out until August 2020, when most establishments, including offices, barbershops, markets, malls, were opened, and unrestricted city movements were allowed. In Uttarakhand, sampling was conducted under partial lockdown conditions, with offices closed and restricted activities permitted in some areas. The observations from RT-qPCR based qualitative genome detection showed that the spread of COVID-19 in North India is highly extensive, and partial lockdowns had no apparent difference in the viral genome load. The Ct values of all three genes (E, RdRp, and N) were found to be in the range of 30-38 (Table 2), which corresponds to a mild to moderate genome load presence inall the wastewater samples. In Jaipur, the Ct values indicate an increase in the genome load of about 10 to 10^3^ fold after the restrictions were lifted. The Ct values in wastewater samples collected from Uttarakhand indicate genome loads within the same orders of the loads present in Jaipur during June 2020. It was the time when restrictions were being lifted partially in the cities. The increase in the viral genome load concurs with the gradual rise in the number of infected individuals, which have risen from 2138 and 720 active cases on first June to 10,260 and 5912 active cases by the end of August 2020 in Jaipur and Haridwar districts respectively (number of cases for Jaipur district obtained from the newspaper, the case numbers for Haridwar as reported by the Department of Medical Health and Family Welfare (https://health.uk.gov.in/)). During the qualitative detection, the presence of gene E was frequently detected with the lowest of Ct values as compared to the other two genes (Genes N and RdRP) (Table 2). In Uttarakhand, the Ct of E gene in four samples out of five positive samples was the lowest compared to those of N and RdRP genes in the same samples. In Jaipur, all the five-times sampling in August 2020 (overall 10 out of 12 samples), showing positive results for the presence of the SARS-CoV-2 genome, had the lowest Ct values for the E gene. Interestingly, for the samples collected initially from Jaipur, Ct values of E and N indicated that the genome loads were still within a difference of 10 times maximum. In contrast, the sample collected later into the time window clearly showed 100 to 1000 times lower genome load reflecting in the Ct values for the E gene than N. Several studies have investigated the role of these viral genes. It has been known that these genes are involved in different steps of viral assembly. Vennemal et al demonstrated that E protein along with M protein alone is independently capable to assemble into viral capsid without requiring any interactions with N or RdRp genes. Self-assembly of SARS-CoV-2 requires interaction of the N gene with viral RNA for compaction and packaging into the viral capsid (Ye et al., 2020). Thus, the observed Ct value trends might be interesting and may indicate towards shedding of viral capsids before the packaging is complete. Alternatively, it could indicate the differential expression rates of these genes in host under different conditions, like, genetic makeup of a community, geographical, climatic, etc. These dynamics in E, N, and RdRP gene detection, therefore might prove useful to understand the viral host interactions and transmission probabilities through wastewater.

A few studies have also tried to correlate the Ct values with genome load and the probability of that load to be infective. Lab level studies were conducted by investigating the percentage of the cell cultures turning positive at various Ct levels of the SARS-CoV-2 genome detected in the clinical samples of sputum and nasal swabs. The study shows that a sample with genome load with Ct values greater or equal to 34 could not infect the cell lines tested and thus postulates that the patients with higher or equal to 34 Ct values may be discharged out (La Scola et al., 2020). However, those studies have been conducted in labs for clinical samples and might not correspond to the infection probabilities that could occur through the contamination in the wastewater. Thus, it would be interesting to investigate the possibilities of transmission routes through contaminated wastewater.

### 3.2. Detection of SARS-CoV-2 RNA in effluent samples corresponds to the role of the biological wastewater treatment process

The presence of the SARS-CoV-2 genome in untreated wastewater is a cause of concern as the wastewater is a potential route of viral transmission to the sanitation workers. Additionally, aerosolization of wastewater during its treatment can promote infection via air provided the viral particle is active. The sludge and treated water from these treatment facilities are used for agricultural purposes, which can put end users’ health at risk. To investigate the probability of such a transmission route, treated wastewater samples from all the sampling sites were collected and checked for the presence of viral RNA.

The samples were collected from both secondary and tertiary treatment stages of wastewater treatment plants. Interestingly, detectable and intact SARS-CoV-2 viral genome was not observed in any treated wastewater samples (Table 3). The sampling was conducted during the lockdown and after lifting of the lockdown regulations. While there is a constant increase in the number of COVID-19 cases, and by extension, the SARS-CoV-2 genome load per sample unit, treated samples were negative for the viral genome presence. It indicates that the wastewater treatment facilities were capable of degrading the viral RNA significantly. The biological treatment stages were capable of removing the intact SARS-CoV-2 genome beyond the detection sensitivity and did not solely depend on the tertiary treatment (i.e., disinfection stage). Direct chlorination of untreated sewage might not show any significant reduction in the detected viral loads if the chlorine demand of the sample is not satisfied. Chlorine demand is directly proportional to the organic waste matter present in the water samples (Rudolf and Ziemba, 1983). Since there is a very large quantity of organic matter in the sewage; it is understandable if chlorination alone is not highly effective. Evidently, Zhang et al. (2020) found an unexpected occurrence of SARS-CoV-2 viral RNA in the septic tank even after disinfection with sodium hypochlorite. They suggested reevaluation of the existing disinfection approach (free chlorine: > 6.5 mg/L after 1.5-hour contact). Thus, highlighting the significant role played by biological treatment systems.

**Table 3.**
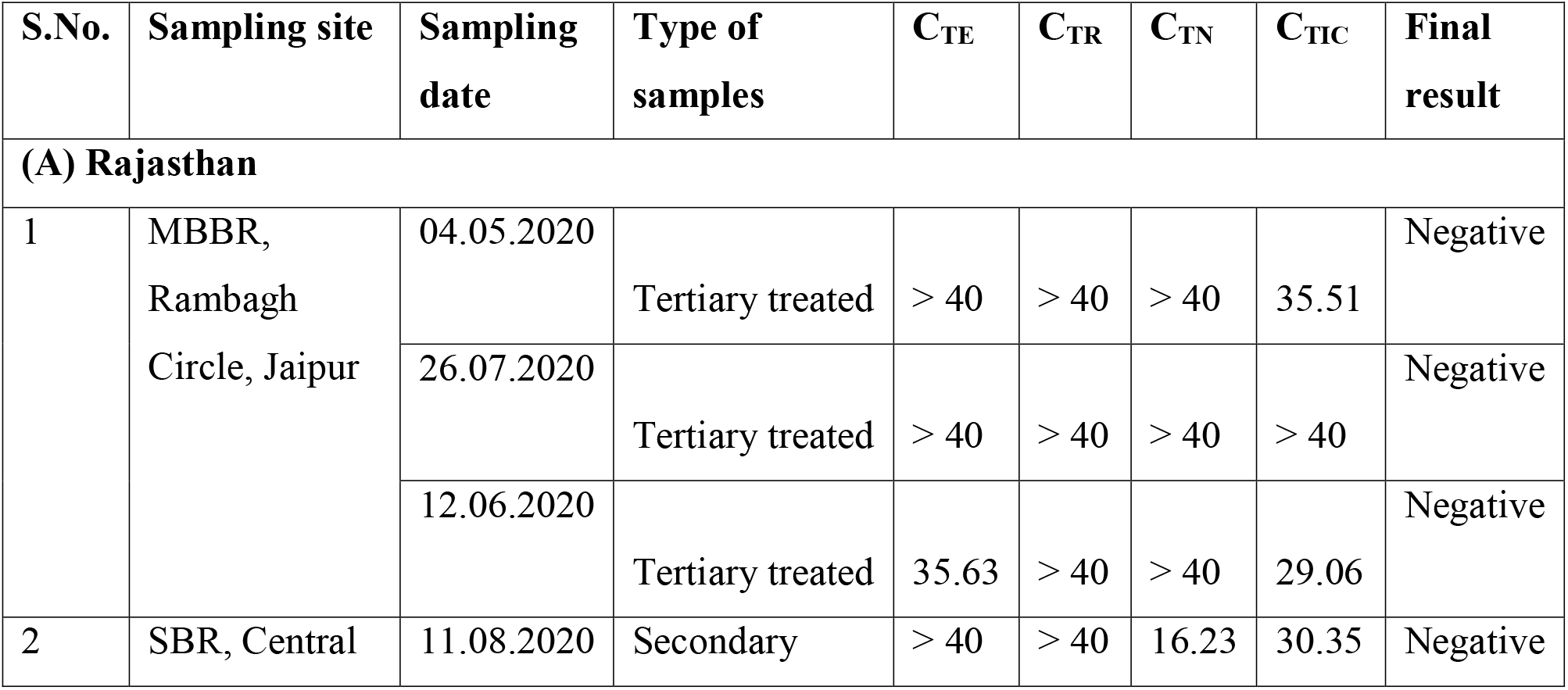
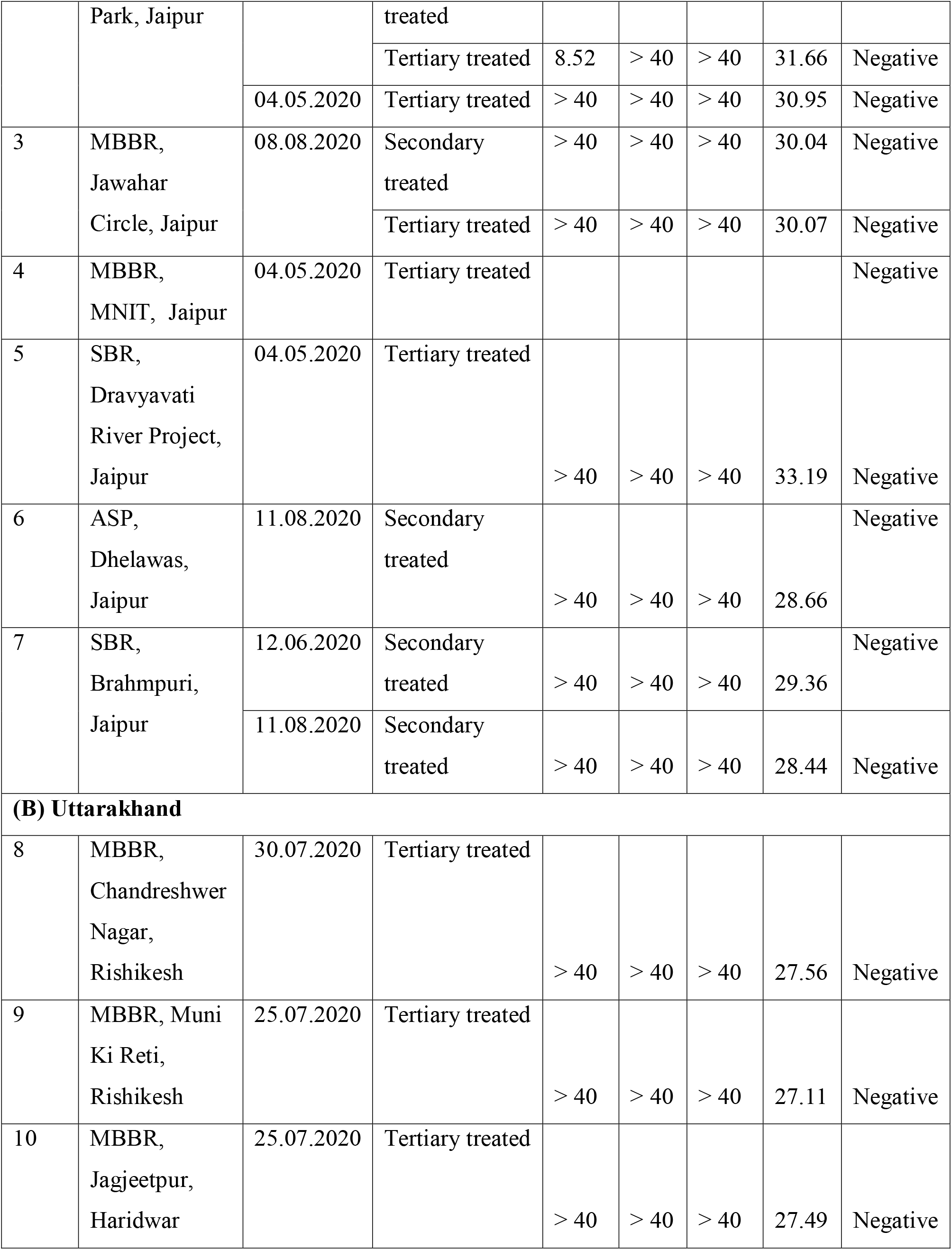
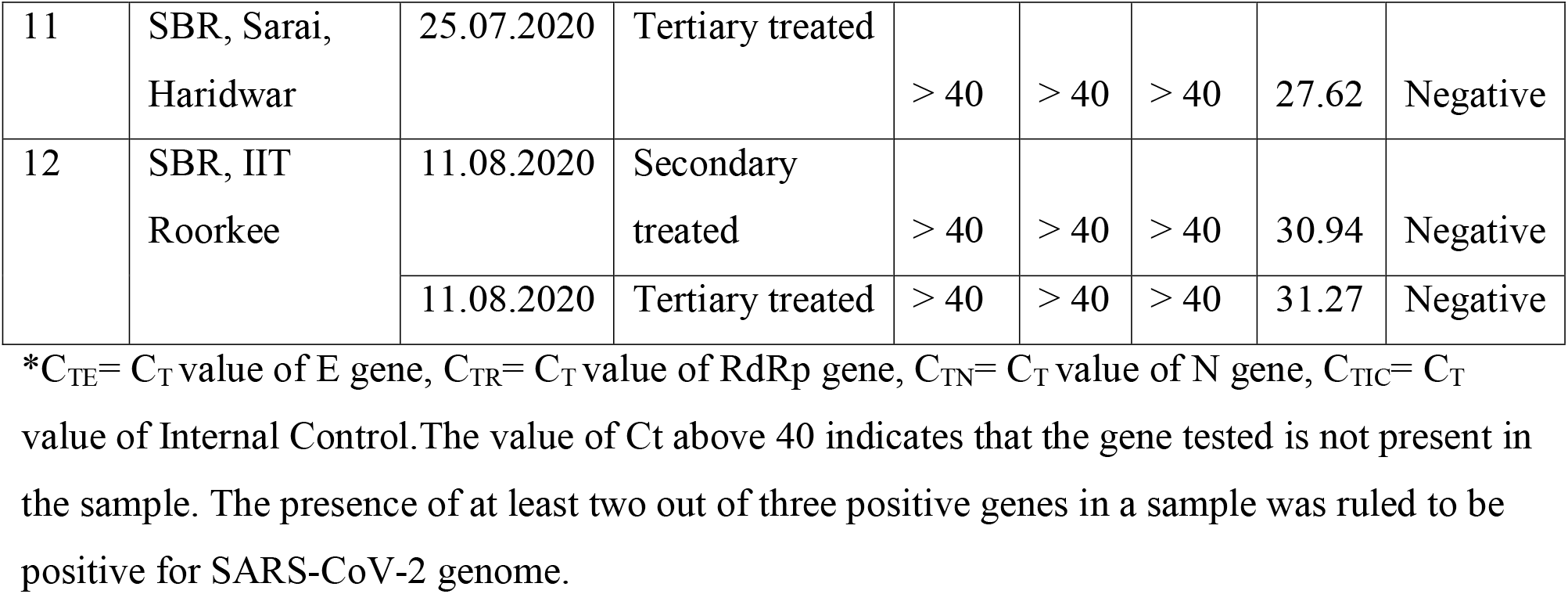
SARS CoV-2 RNA in secondary and tertiary treated wastewater samples collected from WWTPs

## 4. Discussion

This study highlights the effect of an increase in the viral genome load per unit of wastewater. In the post lockdown period (August 2020), the rapid increase in the numbers of COVID-19 patients were corroborated by the decrease in Ct values. Additionally, the genes tested for SARS-CoV-2 in the wastewater showed different gene load levels, as indicated by the Ct values. The E gene seems to be present more abundantly than N and RdRP in the samples. This observation can be explained by either of two reasons. One reason could be the host-pathogen interaction which is different for different populations; thus it is possible that a particular gene is more abundantly or stably expressed in a community. Another reason could be that the E gene is responsible for the structural assembly of the viral particles without interacting with N and RdRp genes (Vennemal et al., 1996) and thus could be simply overexpressed as compared to other SARS-CoV-2 genes in affected individuals leading to shedding of assembled capsids without packaged RNA genome

Throughout the sampling period, the SARS-CoV-2 genome could not be detected in the secondary or tertiary treated samples. However, few studies reported a low probability of viral detection until the tertiary treatment is carried out (Randazzo et al., 2020). The difference in observations could be explained by multiple factors, e.g., the physical factors (temperature, pH), and chemical factors, present as the constituents of the wastewater (i.e., constitutents of wastewater may affect the stability of viral genes.)or the microbiome involved in the biological treatment process itself. The absence of any detectable viral genome in wastewater samples collected post-secondary treatment might indicate the efficacy of the biofilm generated by the microflora in the biological reactors in removing the viral genome loads. This hypothesis is based on several studies that have reported the role of biofilms in the removal of various types of viruses(Quignon et al., 1997; Esfahani et al., 2020). A biofilm can be defined as a well-organized community consisting of cooperating microorganisms immobilized in an extracellular polysaccharide (EPS) matrix (Mortensen, 2014; Maurya, and Raj, 2020). Biofilms can be an association of a single, or multiple species of bacteria, fungi, algae, protozoans, and rotifers in combinations (Maurya and Raj, 2020). These unique combinations allow the matrix to effectively trap and assist in removing the viral particles present in raw wastewater flowing through it. Further studies investigating the composition of the biofilms present in the secondary treatment systems might provide a deep insight into the structures and composition of biofilms, which are specifically effective in SARS-CoV-2 removal.

## 5. Conclusions

Following conclusions can be put forward from this study:

- Ct values corresponding to mild-moderate levels of viral genome loads were observed and direct effects of physical distancing and lockdown regulations on the Ct values could be seen.
- Further, differences in the presence of genes were observed; Ct values of gene E indicated its abundance compared to the N or RdRP gene in the samples collected post lockdown.
- Biological treatment plants might be appropriate to perform with high efficiency in SARS-CoV-2 removal, diminishing any possibility of the fecal route of disease transmission through treated wastewater.
- It also assessed the efficacy of biological treatment followed by tertiary treatment (disinfection) to avoid contamination from treated urban wastewater, and establish a surveillance system through sewage monitoring of the potential virus circulation.
- This study provides a new direction to understand the survival of these viruses in natural conditions, at different temperatures, and in various wastewater types.

## Data Availability

The data in the manuscript is original to the study.

## Acknowledgements

Authors are thankful to the Department of Biotechnology-GoI (GrantNo. BT/RLF/Re-entry/12/2016) for financial support to this research.

## Notes

### Competing Interest Statement

The authors have declared no competing interest.

### Author Declarations

Institutional Ethical clearance obtained.

